# Characteristics of patients with myofascial pain syndrome of the low back

**DOI:** 10.1101/2023.10.15.23297051

**Authors:** Pao-Feng Tsai, Joseph L. Edison, Chih-Hsuan Wang, Michael W. Gramlich, Kailea Manning, Gopikrishna Deshpande, Adil Bashir, JoEllen Soften

**Author notes:** Corresponding Author: College of Nursing, Auburn University, 710 S Donahue Dr, Auburn, AL 36849.

## Abstract

**Objective:** Identify characteristics of patients with myofascial pain syndrome (MPS) of the low back.

**Methods:** Twenty-five subjects with myofascial trigger point(s) [MTrP(s)] on the low back participated in this cross-sectional study. The location, number and type (active or latent) of MTrPs were verified by ultrasound; additionally, data on pain pressure threshold, physical function, quality of life, disability, pain catastrophizing, pain self-efficacy, kinesiophobia, emotional health, exercise motivation and pain were collected. Descriptive statistics, Chi-square, one-way ANOVAs and factorial ANCOVA were used to achieve study objectives.

**Results:** No statistically significant differences in variables were found between types of MTrPs: Group 1 (Active, n=13), 2 (Latent, n=5) 3 (Atypical, no twitching but with spontaneous pain, n=2) and 4 (Atypical, no twitching and no spontaneous pain, n=5) except the number of MTrPs, current pain, and worst pain (p=.01-.001). There were interaction effects between spontaneous pain and twitching response on reports of physical function, current pain level, and worst pain level (p=.04-.002). Participants in Group 3 reported lower levels of physical function, higher levels of current pain, and higher levels of worst pain compared to those in Group 4. Participants in Group 1 and 2 had similar levels of physical function, current pain and worst pain.

**Discussion:** Number of MTrPs is most closely associated with the level of pain. Spontaneous pain report seems to be a decisive factor associated with poor physical function; however, twitching response is not.

## Introduction

Myofascial pain syndrome (MPS) is a regional pain disorder that causes small nodules of tight tissue called myofascial trigger points (MTrPs). Patients with active MTrPs typically present with spontaneous and recognizable pain. Latent MTrPs may produce local or referred pain after palpation and demonstrate myofascial dysfunction.^1^ It has been hypothesized that mechanical factors, such as poor posture, abnormal gait and prior injury, result in muscle overload and eventually lead to muscle contraction and formation of MTrPs and pain reaction.^2, 3^ Alternative theories postulated that psychological factors, such as stress, are major elements contributing to the activation and maintenance of MTrPs and play a significant role in the intensity of the perception of pain.^3^ This stems from the observation that myofascial pain is referred pain and hence, central sensitization may have a role to play in that process.^4^

Diagnosis of MPS currently depends on physical examination and patient’s self-report of pain. Lack of clinician training and skill, inconsistent diagnosis criteria and terminology, and subjectivity of diagnosing may result in ignoring patient symptoms and misdiagnosis ^5, 6, 7^ In addition, patients may not feel that it is necessary or important to report symptoms due to the chronic nature of the pain. Therefore, the prevalence rate of MPS is unclear. It is estimated that 30% of patients visiting primary care clinics and 85% visiting pain clinics suffer from MPS. Approximately 30% - 93% of patients with widespread pain also present with MTrPs.^1, 8^ Specifically, studies found that 63.5%-90% of patients with low back pain suffered from MPS.^9, 10^ Like most chronic pain conditions, MPS patients are likely under treated.^11^

A few small studies have investigated the characteristics of patients with MPS of the low back as well as characteristics of MTrPs on the low back. Patient demographics seem similar to patients with non-specific low back pain.^9^ A study comparing low back pain and healthy adults showed that Individual patients showed multiple MTrPs on the low back^12^ and the intensity of their low back pain episode was associated with the number of active MTrPs measured.^12^ This study neither confirmed the presence of trigger points using an ultrasound procedure nor compared the characteristics of patients with active and latent trigger points.

Therefore, the aim of this pilot study was to identify characteristics of patients with MPS of the low back and compare the difference in characteristics between patients with active and latent MTrPs. Ultrasound was utilized to verify the existence of MTrPs. This study included qualitative and quantitative components at the individual level as well as ultrasound evaluation at the MTrP level. This study reports the individual level quantitative components of the larger study.

## Materials and Methods

### Sample

This study was part of a larger cross-sectional study that investigated qualitative and quantitative components of MTrPs at the individual level and also ultrasound evaluation at the MTrP level. The current study reports the individual level quantitative components of the individuals with MPS. This project was IRB approved from author’s institutions. It was conducted between 8-30-2021 and 6-30-2022. Inclusion criteria included participants who (1) self-reported having chronic low back pain with palpable MTrP(s) on the low back muscles and hypo-echogenicity of MTrP region(s) confirmed by ultrasound examination, (2) were English-speaking, and (3) were ambulatory without a cane or walker. Exclusion criteria included having (1) major illness, such as cancer, (2) major surgery within 6 months, (3) major psychiatric disorder, such as bipolar disorder and depression, (4) cognitive impairment or (5) other painful conditions of the low back. Participants were recruited from the local community using a snowball sampling procedure.

### Identifying location, number and type of MTrPs

To screen for MPS, the participant was positioned prone on an examination table. To identify the site and number of MTrPs, the participant was examined by an osteopathic doctor using physical examination and palpation to determine the presence or absence of MTrP(s). The osteopathic doctor regularly conducts this examination as part of their clinical practice. A total of 25 participants was screened and all were identifying having MTrPs. As this study is of a pilot nature, only three MTrPs were randomly selected for each participant for further evaluation, and an ultrasound procedure was applied to confirm the palpation findings of the three MTrPs. A latent MTrP is defined as a tender spot within a taut band of a skeletal muscle, which is clinically associated with a local twitch response, tenderness and/or referred pain upon manual examination. Active MTrPs are similar to latent MTrPs but show spontaneous local or referred pain.^3^ If nodules were readily visualized under ultrasound-guided examination, participants with a spontaneous pain complaint but no twitching response when examined, or no spontaneous pain and no twitching response were categorized as having atypical MTrPs. Participants were divided into four groups for analysis:

1. Group 1 (active MTrP group): Participants had at least one active MTrP.
2. Group 2 (latent MTrP group): Participants had no active MTrPs but had at least one latent MTrP.
3. Group 3 (atypical MTrP group): Participants had no active and no latent MTrP but at least one nodule on the low back muscle was visible in the ultrasound exam and had spontaneous pain.
4. Group 4 (atypical MTrP group): Participants had no active and no latent MTrP but at least one nodule on the low back muscle was visible in the ultrasound exam and had no spontaneous pain.

### Measurements

#### Demographic questionnaire

A structured interview was conducted to collect patient demographic information. Items such as date of birth, sex, ethnicity, race, marital status, and highest level of education completed were adopted from the National Healthy Worksite Demographic Questions Measure.^13^ Participant’s employment status was adopted from the 2021 Behavioral Risk Factor Surveillance System Questionnaire.^14^

#### Pain Pressure Threshold (PPT)

PPT was used to objectively measure participant pain for each MTrP.^15^ The Wagner Algometer FPX 25 (Wagner Instruments, Greenwich, CT) was utilized to apply and measure pressure. Patients were instructed to say “Stop” when they felt pain in pounds of force (lbf). PPT was measured three times at each MTrP. The three measurements were averaged and utilized for the PPT score for each MTrP.

#### Five Repetition-Sit to Stand Test (5R-STS)

(5R-STS) is a reliable,^16^ clinical assessment used to objectively test movements used for everyday life (i.e., sitting down and standing up). Beginning in a sitting position, participants were instructed to stand up fully and sit down firmly on the seat five times as fast as possible on the command “go.” Researchers recorded the time using a stopwatch from the time “go” was announced to the time the participant sat down for the fifth time.^17^ ^16^

#### Timed Up and Go (TUG)

The TUG assessment is a quick, objective measurement to assess physical function n that demonstrates good test-retest reliability.^18^ The assessment begins with the participant sitting in a chair. On the command “go,” the participant is to walk three meters at a regular pace, turn around, and walk back to the chair, returning to a sitting position. The staff recorded the time it took to complete this task using a stopwatch. ^18^

#### Short form health outcomes survey (SF-20)

The SF-20 is a twenty-item measure that is made up of six health concepts including physical functioning (6 items), role functioning (2 items), social functioning (1), mental health (5), health perceptions (5), and pain (1). Each response item is assigned a value of one to five. Scoring is achieved by changing the raw score. Reliability coefficients range from 0.81 - 0.88,^19^ which are similar to the full-length versions. Cronbach alpha reliability for the current study ranged between 0.75-0.93, except the mental health (0.47).

#### Oswestry Disability Index (ODI)

The ODI evaluates disability due to low back pain. It consists of ten items to assess pain intensity, personal care, lifting, walking, sitting, standing, sleeping, sex life, social life, and traveling.^20^ Each response item is assigned a value of 0-5. The scoring is then conducted by adding up each item’s score and then dividing that total number by the total amount possible and multiplying by 100. A lower percentage represents a better low back health score whereas a higher percentage represents worse back health. The ODI‘s Cronbach’s Alpha reliability within adult populations were 0.76 - 0.88.^21, 22^ Cronbach alpha reliability for the current study was 0.74.

#### Pain Catastrophizing Scale (PCS)

The PCS assesses catastrophic thinking regarding pain.^23^ This 13 item self-report scale includes three subscales: rumination, magnification and helplessness. The items are rated on a 5-point scale ranging from 0 (not at all) to 4 (all the time). Scoring consists of summing all the item scores to calculate a total score, which can range from 0-52. A higher score is associated with more pain catastrophizing. Cronbach’s alpha coefficients were 0.89 - 0.95.^24, 25^ Cronbach alpha reliability for the current study was 0.92.

#### Pain Self-Efficacy Questionnaire (PSEQ)

The PSEQ asks participants to rate how confident they are in performing various activities, despite their current pain. It is a 10-item measure and responses are made on a 7-point scale ranging from 0 (not at all confident) to 6 (completely confident). The scale is scored by summing all the items to produce a total score ranging from 0-60, where higher scores indicate higher levels of self-efficacy beliefs. This measure has previously been found to be reliable with a Cronbach’s alpha coefficient of .94.^26^ The Cronbach alpha reliability for the current study was .90.

#### Tampa Scale of Kinesiophobia (TSK)

<u>The TSK</u> assessed participant’s fear for movement and/or re-injury.^27^ The measure includes 17-items using a 4-point Likert-type scale ranging from 1 (strongly disagree) to 4 (strongly agree). Four items are inversely scored. After these scores have been transformed, the item scores are added together so that a higher score suggests more fear. The reliability for this scale has been shown with a Cronbach’s alpha of 0.76.^27^ The Cronbach alpha reliability for the current study was 0.76.

#### The Depression, Anxiety, and Stress Scale (DASS-21)

<u>The DASS-21</u> is a self-reported measure to assess the emotional states of depression, anxiety, and stress.^28^ It consists of 21 items with seven items belonging to each subscale (Stress, Anxiety and Depression). A 4-point response scale is used ranging from 0 (did not apply to me at all) to 3 (applied to me very much or most of the time). Scoring is conducted by adding up each item belonging to the three different subscales and multiplying by 2 for each final score. In previous work, Cronbach’s alpha coefficients for each subscale were between 0.82 and 0.94.^28, 29^ Cronbach alpha reliability coefficients for the current study were 0.77, 0.70 and 0.45 for Stress, Anxiety and Depression subscales, respectively.

#### Exercise Motivation Inventory-2 (EMI-2)^30^

The EMI-2 is a 51-item, self-report measure designed to assess motives for exercising.^31^ The measure includes a 5-point response scale where 0 indicates “Not at all true for me” and 5 indicates “Very true for me.” Scoring is achieved by adding the respective items and then finding the mean of these totals for each subscale. The inventory has 14 subscales, including Stress Management, Revitalization, Enjoyment, Challenge, Social Recognition, Affiliation, Competition, Health Pressures, Health Avoidance, Positive Health, Weight Management, Appearance, Strength & Endurance, and Nimbleness. Cronbach’s alpha coefficients for each subscale have been found to be between 0.66 and 0.95.^31, 32^ Cronbach alpha reliability coefficients for the current study were equal to or above 0.80 except for Social Recognition (0.78) and Health Pressures (0.64) subscales.

#### The Body Mass Index

The Body Mass Index (BMI) was determined by participants height and weight, using the formula: weight (lb) / [height (in)]^2^ x 703.^33^

#### Pain

Three items were adopted from Brief Pain Inventory^34^ including patient reports of their current low back pain, best pain level (i.e., lowest pain rating) and the worst pain level (i.e., highest pain rating) they have had in the past 24 hours. Using the Numeric Pain Rating Scale^35^, a visual scale ranging from 0 (no pain) to 10 (worst pain imaginable), participants were asked to rate each of these pain levels regarding the pain in their low back.

#### Data management and analysis

After completing the data collection, the project manager entered the de-identified data into Excel worksheets. The project statistician evaluated if the data was entered correctly and made corrections if needed. The project statistician then imported the Excel worksheets into the Statistics Package for the Social Sciences for Windows version v28.0 (IBM Corp., Armonk, N.Y., USA) for data analysis.

A series of Chi-square Independence Tests was used to examine the distribution of each variable among different groups, such as gender, ethnicity, educational level, etc. A series of one-way ANOVAs was conducted to examine the mean differences of each continuous demographic variable as well as the mean scores of subscales among the four groups, such as age, BMI, the level of tenderness, etc. Finally, a series of Factorial ANCOVAs, using age, gender, and ethnicity (dummy coded) as the covariate, were conducted to examine if there were twitching or pain main effects, and the interaction effect on each scale. Frequencies were used to describe treatment and strategies used to manage MPS.

## Results

### Descriptive results

Twenty-five participants with low back pain participated in this pilot study, reporting active (n=13), latent (n=5), and atypical MTrPs (n=7). Table 1 describes the demographic characteristics and general health information of the study sample and subgroups. with no significant differences between groups found. Seventy-two percent were women (*n*=18), the majority identified as White (64%, *n*=16) and reported being employed (60%, *n*=15), all participants had at least a college degree or higher, and the mean age was 34.56 (SD=10.75). The majority rated their health as good and above (92%, *n*=23) and only thirty-two percent of participants had pain at the moderate level in the last four weeks. Eighty percent of participants (80%, *n*=20) reported that they took pain medication for MPS, mostly over the counter medication. The mean BMI was 26.84 (*SD*=5.45) which is considered average, and the mean number of self-reported current medical and surgical conditions was 1.52 (*SD*=1.29). The mean number of MTrPs identified on the low back and duration of MPS was 4.36 (*SD*=0.91) and 82.80 months (*SD*= 85.73 months), respectively.

**Table 1.** Demographic Characteristics of the Study Sample (*N*=25)

| Variables | Total<br>N=25 |  | Group 1<br>n=13 |  | Group 2<br>n=5 |  | Group 3<br>n=2 |  | Group 4<br>n=5 |  | p value |
| --- | --- | --- | --- | --- | --- | --- | --- | --- | --- | --- | --- |
|  | n | % | n | % | n | % | n | % | n | % |  |
| Sex | | | | | | | | | | | $\chi^2(3)=3.79, p=.29$ |
| Male | 7 | 28.0 | 5 | 38.5 | 0 | 0.0 | 0 | 0.0 | 2 | 40.0 |  |
| Female | 18 | 72.0 | 8 | 61.5 | 5 | 100.0 | 2 | 100.0 | 3 | 60.0 |  |
| Ethnicity | | | | | | | | | | | $\chi^2(6)=2.91, p=.82$ |
| White | 16 | 64.0 | 9 | 69.2 | 3 | 60.0 | 1 | 50.0 | 3 | 60.0 |  |
| Black/African American | 3 | 12.0 | 2 | 15.4 | 1 | 20.0 | 0 | 0.0 | 0 | 0.0 |  |
| Asian | 6 | 24.0 | 2 | 15.4 | 1 | 20.0 | 1 | 50.0 | 2 | 40.0 |  |
| Employment | | | | | | | | | | | $\chi^2(3)=1.76, p=.62$ |
| Employed | 15 | 60.0 | 8 | 61.5 | 4 | 80.0 | 1 | 50.0 | 2 | 40.0 |  |
| Unemployed/Students | 10 | 40.0 | 5 | 38.5 | 1 | 20.0 | 1 | 50.0 | 3 | 60.0 |  |
| Education | | | | | | | | | | | $\chi^2(3)=0.96, p=.81$ |
| Some college | 1 | 4.0 | 1 | 7.7 | 0 | 0.0 | 0 | 0.0 | 0 | 0.0 |  |
| College graduate | 24 | 96.0 | 12 | 92.3 | 5 | 100.0 | 2 | 100.0 | 5 | 100.0 |  |
| Marital status | | | | | | | | | | | $\chi^2(6)=4.42, p=.62$ |
| Married | 18 | 72.0 | 10 | 76.9 | 3 | 60.0 | 2 | 100.0 | 3 | 60.0 |  |
| Unmarried couple | 3 | 12.0 | 2 | 15.4 | 1 | 20.0 | 0 | 0.0 | 0 | 0.0 |  |
| Never married | 4 | 16.0 | 1 | 7.7 | 1 | 20.0 | 0 | 0.0 | 2 | 40.0 |  |
| Health | | | | | | | | | | | $\chi^2(9)=9.74, p=.37$ |
| Excellent | 3 | 12.0 | 2 | 15.4 | 0 | 0.0 | 0 | 0.0 | 1 | 20.0 |  |
| Very Good | 9 | 36.0 | 5 | 38.5 | 2 | 40.0 | 0 | 0.0 | 2 | 40.0 |  |
| Good | 11 | 44.0 | 6 | 46.2 | 3 | 60.0 | 1 | 50.0 | 1 | 20.0 |  |
| Fair | 2 | 8.0 | 0 | 0.0 | 0 | 0.0 | 1 | 50.0 | 1 | 20.0 |  |
| Pain in past 4 weeks | | | | | | | | | | | $\chi^2(6)=8.99, p=.17$ |
| Very Mild | 7 | 28.0 | 2 | 15.4 | 1 | 20.0 | 0 | 0.0 | 4 | 80.0 |  |
| Mild | 10 | 40.0 | 6 | 46.2 | 2 | 40.0 | 1 | 50.0 | 1 | 20.0 |  |
| Moderate | 8 | 32.0 | 5 | 38.5 | 2 | 40.0 | 1 | 50.0 | 0 | 0.0 |  |
| Pain medication for MPS | | | | | | | | | | | $\chi^2(3)=6.73, p=.08$ |
| Yes | 20 | 80.0 | 12 | 92.3 | 4 | 80.0 | 2 | 100.0 | 2 | 40.0 |  |
| No | 5 | 20.0 | 1 | 7.7 | 1 | 20.0 | 0 | 0.0 | 3 | 60.0 |  |
| Mean age (SD) | 34.56 (10.75) | | 34.0 (11.71) | | 34.80 (7.29) | | 34.50 (17.68) | | 35.60 (12.18) | | $F(3,21)=0.02, p=1.00$ |
| Mean BMI (SD) | 26.84 (5.45) | | 28.68 (6.32) | | 24.45 (3.31) | | 27.59 (3.66) | | 24.17 (4.12) | | $F(3,21)=1.26, p=.31$ |
| Mean numbers of current health conditions (SD) | 1.52 (1.29) | | 1.42 (1.38) | | 1.67 (2.08) | | 2.50 (0.71) | | 1.25 (0.50) | | $F(3,21)=0.44, p=.73$ |
| Average pain duration in month (SD) | 82.8 (85.73) | | 60.46 (75.74) | | 120.00 (83.57) | | 36.00 (33.94) | | 122.4 (114.91) | | $F(3,21)=1.19, p=.34$ |

Overall, the mean number of MTrPs found on the low back was 4.36 (*SD*=0.91). The mean reported pain level of MTrPs on a numeric pain rating scale of 0-10, when palpated, was 3.97 (*SD*=1.43), but the MTrP with the most severe pain level had a mean of 5.44 (*SD*=1.47). The mean pressure pain threshold on MTrPs was 4.43 lbf (*SD*=2.23) and the most sensitive MTrPs only tolerated 3.63 lbf (*SD*=1.89).

Objective measure of physical function, STS, was 10.60 (*SD*=2.48) seconds, indicating that it took the study sample longer to complete the exercise compared to healthy population.^36^ Similarly, the mean of TUG was 9.36 (SD=1.76) seconds. This result showed that it took longer for the study participants as a whole to complete the exercise even compared to a population of older adults.^37^

The physical health component of quality of life was 83.34% (*SD*=18.94%), role functioning was 88.00% (*SD*=25.12%), social functioning was 92.80% (*SD*=11.37%), mental health was 78.48% (SD=9.98%), health perceptions was 80.28% (SD=12.37%), and the pain component was 40.80% (*SD*=15.80%). The participants reported a better quality of life in all components when compared with a sample of similarly aged participants who have been diagnosed with low back pain.^38^ For the ODI score measuring disability, participants present a slightly higher score (*M*=11.44, *SD*=7.8) compared to a healthy, but scored exceedingly lower than a population of patients with chronic back pain.^20^ The PCS score for the study sample suggests participants experience moderate pain catastrophizing (*M*=11.48, *SD*=9.75), similar to a population with chronic low back pain.^39, 40^ The participant’s mean PSEQ score (*M*=53.52*, SD*=6.63) suggests they have greater pain self-efficacy beliefs (i.e., they are much more confident in their ability to handle pain) compared to a mean population of individuals with low back pain.^41^ The mean score on the TSK for the study’s sample (*M*=32.60, *SD*=6.34) falls below the cut off score of 37 for chronic low back pain, suggesting the study sample as a whole are less fearful of movement or reinjury.^42^ Stress, anxiety and depression scores of the emotional health measure (DASS 21) were 7.92 (*SD*=6.84), 3.44 (*SD*=5.43) and 1.76 (*SD*=2.11), respectively meaning on average participants reported a normal level of stress, anxiety, and depression.^43^ The mean exercise motivation inventory (EMI) subscales were between 1.00-4.24, meaning participants in the study had similar levels of motivation for exercising compared with a healthy sample of younger and middle-aged adults.^44^ Current pain, best pain and worst pain levels were 2.96 (*SD*=1.95), 1.04 (*SD*=1.40), and 5.00 (*SD*=2.57) respectively, indicating that participants’ current and best pain were categorized as “mild” while their worst reported pain was considered to be in the “moderate” cutoff.^35^

### Difference between patients in 4 groups

Results indicate no statistically significant differences in the four groups in clinical assessment and questionnaire intakes, with the exception of the number of trigger points and verbal report of pain (Table 2). Participants in pain without twitching (Group 3), had more trigger points than those in the group who had no pain and no twitching (Group 4) with a significant effect size, *F*(3.21)=3.83, *p*=.03, η^2^=.35. In addition, participants from different groups reported different levels of current pain and worst pain in the past 24 hours with a large effect size, *F*(3,21)=5.42, *p*=.01, η^2^=.44, *F*(3,21)=7.71, *p*=.001, η^2^=.52, respectively. The Bonferroni post hoc test results indicated that participants with pain (Group 1 [active MTrP] and Group 3 [atypical MTrP]) reported significantly higher levels of current pain compared with the participants in Group 4, *p*=.04, *p*=.01, respectively. Participants in Group 1 and Group 3 also reported higher levels of worst pain in the past 24 hours compared with participants in Group 4, *p*<.001, *p*=.03, respectively.

**Table 2.** Results of Clinical Assessment and Questionnaire Intakes by Groups (*N*=25)

| Scale | Total |  | G1 (n=13) |  | G2 (n=5) |  | G3 (n=2) |  | G4 (n=5) |  | F<br>(3,21) | p | η² | Observed<br>Power |
| --- | --- | --- | --- | --- | --- | --- | --- | --- | --- | --- | --- | --- | --- | --- |
|  | M | SD | M | SD | M | SD | M | SD | M | SD |  |  |  |  |
| MTrPs |  |  |  |  |  |  |  |  |  |  |  |  |  |  |
| Number | 4.36 | 0.91 | 4.62 | 0.77 | 4.00 | 0.71 | 5.50 | 0.71 | 3.60 | 0.89 | 3.83 | .03* | .35 | .74 |
| Duration | 82.80 | 85.73 | 60.46 | 75.74 | 120.00 | 83.57 | 36.00 | 33.94 | 122.40 | 114.91 | 1.19 | .34 | .15 | .27 |
| Tenderness (Average) | 3.97 | 1.43 | 4.30 | 1.61 | 3.46 | 0.84 | 5.00 | 1.88 | 3.20 | 0.96 | 1.33 | .29 | .16 | .30 |
| Tenderness (Severe) | 5.44 | 1.47 | 5.69 | 1.49 | 5.00 | 1.58 | 6.50 | 2.12 | 4.80 | 1.10 | 0.93 | .45 | .12 | .22 |
| PPT (Average) | 4.43 | 2.23 | 4.12 | 2.61 | 4.21 | 2.05 | 3.50 | 0.90 | 5.79 | 1.33 | 0.82 | .50 | .11 | .20 |
| PPT (Severe) | 3.63 | 1.89 | 3.33 | 2.29 | 3.42 | 1.34 | 3.20 | 0.71 | 4.79 | 1.29 | 0.77 | .53 | .10 | .19 |
| STS | 10.60 | 2.48 | 10.30 | 2.46 | 11.86 | 3.27 | 10.99 | 0.78 | 9.96 | 2.25 | 0.59 | .63 | .08 | .15 |
| TUG | 9.36 | 1.76 | 9.49 | 1.90 | 9.32 | 1.16 | 10.96 | 1.86 | 8.43 | 1.80 | 1.04 | .40 | .13 | .24 |
| SF-20 (%) |  |  |  |  |  |  |  |  |  |  |  |  |  |  |
| Physical Function | 83.34 | 18.94 | 85.26 | 16.37 | 73.34 | 27.25 | 62.50 | 5.94 | 96.68 | 4.55 | 2.56 | .08 | .27 | .55 |
| Role Function | 88.00 | 25.12 | 88.46 | 29.96 | 85.00 | 22.36 | 75.00 | 35.36 | 95.00 | 11.18 | 0.30 | .82 | .04 | .10 |
| Social Function | 92.80 | 11.37 | 92.31 | 13.01 | 88.00 | 10.95 | 90.00 | 14.14 | 100.00 | 0.00 | 1.02 | .41 | .13 | .24 |
| Mental Health | 78.48 | 9.98 | 76.62 | 11.18 | 78.02 | 9.12 | 82.00 | 2.83 | 82.40 | 10.04 | 0.46 | .71 | .06 | .13 |
| Health Perceptions | 80.28 | 12.37 | 81.47 | 11.16 | 75.96 | 15.38 | 66.60 | 5.09 | 86.16 | 12.71 | 1.25 | .32 | .15 | .29 |
| Pain | 40.80 | 15.79 | 44.62 | 14.50 | 44.0 | 16.73 | 50.00 | 14.14 | 26.00 | 8.94 | 3.06 | .05 | .30 | .63 |
| ODI | 11.44 | 7.8 | 14.00 | 8.12 | 13.20 | 6.72 | 10.00 | 2.83 | 3.60 | 4.10 | 2.75 | .07 | .28 | .58 |
| PCS | 11.48 | 9.75 | 8.15 | 5.21 | 15.60 | 9.63 | 8.00 | 8.49 | 17.40 | 16.46 | 1.62 | .22 | .19 | .36 |
| PSEQ | 53.52 | 6.63 | 53.85 | 5.16 | 48.40 | 10.41 | 58.00 | 1.41 | 56.00 | 4.95 | 1.67 | .20 | .19 | .37 |
| TSK | 32.60 | 6.34 | 31.15 | 5.97 | 36.80 | 8.70 | 36.00 | 1.41 | 30.80 | 4.38 | 1.34 | .29 | .16 | .30 |
| DASS 21 |  |  |  |  |  |  |  |  |  |  |  |  |  |  |
| Stress | 7.92 | 6.84 | 9.38 | 5.38 | 4.00 | 4.00 | 4.00 | 5.66 | 9.60 | 11.52 | 1.07 | .38 | .13 | .25 |
| Anxiety | 3.44 | 5.43 | 3.23 | 4.87 | 1.20 | 1.79 | 6.00 | 8.49 | 5.20 | 8.44 | 0.58 | .63 | .08 | .15 |
| Depression | 1.76 | 2.11 | 1.54 | 1.45 | 0.40 | 0.89 | 3.00 | 4.24 | 3.20 | 3.03 | 1.96 | .15 | .22 | .43 |
| EMI |  |  |  |  |  |  |  |  |  |  |  |  |  |  |
| Stress Management | 3.13 | 1.63 | 3.02 | 1.56 | 3.40 | 1.57 | 3.63 | 0.53 | 2.95 | 2.40 | 0.13 | .94 | .02 | .07 |
| Revitalization | 2.88 | 1.62 | 2.79 | 1.56 | 3.00 | 1.72 | 3.17 | 0.24 | 2.87 | 2.34 | 0.04 | .99 | .01 | .06 |
| Enjoyment | 2.51 | 1.86 | 2.35 | 1.74 | 2.90 | 2.19 | 2.25 | 1.06 | 2.65 | 2.52 | 0.11 | .95 | .02 | .07 |
| Challenge | 2.08 | 1.50 | 2.27 | 1.62 | 1.80 | 1.24 | 2.00 | 1.06 | 1.90 | 1.88 | 0.14 | .94 | .02 | .07 |
| Social Recognition | 1.00 | 0.93 | 1.06 | 1.05 | 1.25 | 0.95 | 0.50 | 0.71 | 0.80 | 0.78 | 0.37 | .77 | .05 | .11 |
| Affiliation | 1.49 | 1.29 | 1.02 | 0.96 | 2.05 | 1.57 | 2.13 | 1.59 | 1.90 | 1.60 | 1.27 | .31 | .15 | .29 |
| Competition | 1.58 | 1.61 | 1.56 | 1.52 | 1.65 | 1.90 | 1.00 | 0.71 | 1.80 | 2.15 | 0.11 | .95 | .02 | .07 |
| Health Pressures | 2.03 | 1.43 | 1.90 | 1.33 | 2.20 | 1.28 | 2.33 | 0.94 | 2.07 | 2.23 | 0.08 | .97 | .01 | .06 |
| Health Avoidance | 3.83 | 1.08 | 3.79 | 0.84 | 3.87 | 1.12 | 4.33 | 0.47 | 3.67 | 1.84 | 0.17 | .92 | .02 | .08 |
| Positive Health | 4.24 | 0.94 | 4.31 | 0.60 | 4.27 | 0.98 | 4.83 | 0.24 | 3.80 | 1.66 | 0.63 | .60 | .08 | .16 |
| Weight Management | 3.99 | 1.32 | 3.56 | 1.64 | 4.20 | 0.96 | 4.50 | 0.00 | 4.70 | 0.33 | 1.10 | .37 | .14 | .25 |
| Appearance | 3.12 | 1.22 | 2.69 | 1.42 | 3.25 | 0.68 | 3.38 | 0.18 | 4.00 | 0.85 | 1.56 | .23 | .18 | .35 |
| Strength & Endurance | 3.66 | 1.33 | 3.94 | 0.79 | 3.65 | 1.42 | 3.13 | 1.59 | 3.15 | 2.32 | 0.51 | .68 | .07 | .14 |
| Nimbleness | 3.15 | 1.31 | 3.23 | 1.05 | 3.33 | 0.53 | 3.17 | 0.24 | 2.73 | 2.52 | 0.20 | .90 | .03 | .08 |
| Numeric Pain Rating |  |  |  |  |  |  |  |  |  |  |  |  |  |  |
| Current Pain | 2.96 | 1.95 | 3.69 | 1.89 | 2.80 | 1.10 | 4.50 | 0.71 | 0.60 | 0.89 | 5.42 | .01* | .44 | .88 |
| Best Pain past 24 Hr | 1.04 | 1.40 | 1.23 | 1.59 | 0.80 | 1.30 | 2.00 | 1.41 | 0.40 | 0.89 | 0.77 | .52 | .10 | .19 |
| Worst Pain past 24 Hr | 5.00 | 2.57 | 6.23 | 1.83 | 4.60 | 2.70 | 6.50 | 0.71 | 1.60 | 1.14 | 7.71 | .001** | .52 | .97 |

### Pain report, twitching response, and interaction on patients reported outcomes

After controlling the covariates including age, gender and race, results indicated there were interaction effects between pain report and twitching response on participants reports of physical function, current pain level, and worst pain level in the past 24 hours with a large effect size, *F*(1,17)=5.10, *p*=.04, η^2^=.23; *F*(1,17)=13.03, *p*=.002, η^2^=.43; *F*(1,17)=4.79, *p*=.04, η^2^=.22, respectively (Table 3 and Table 4). Participants who experienced spontaneous pain (Orange line) but no twitching (G3) reported lower levels of physical function (Figure 1), higher levels of current pain (Figure 2), and higher levels of worst pain (Figure 3) in the past 24 hours compared to those who had both spontaneous pain and twitching response (G1). However, participants who did not have spontaneous pain reported an opposite pattern (Blue line). They reported higher levels of physical function, lower levels of current pain and lower levels of worst pain in the past 24 hours if they had no twitching (G4) response compared to those who had twitching response (G2).

**Figure 1.**
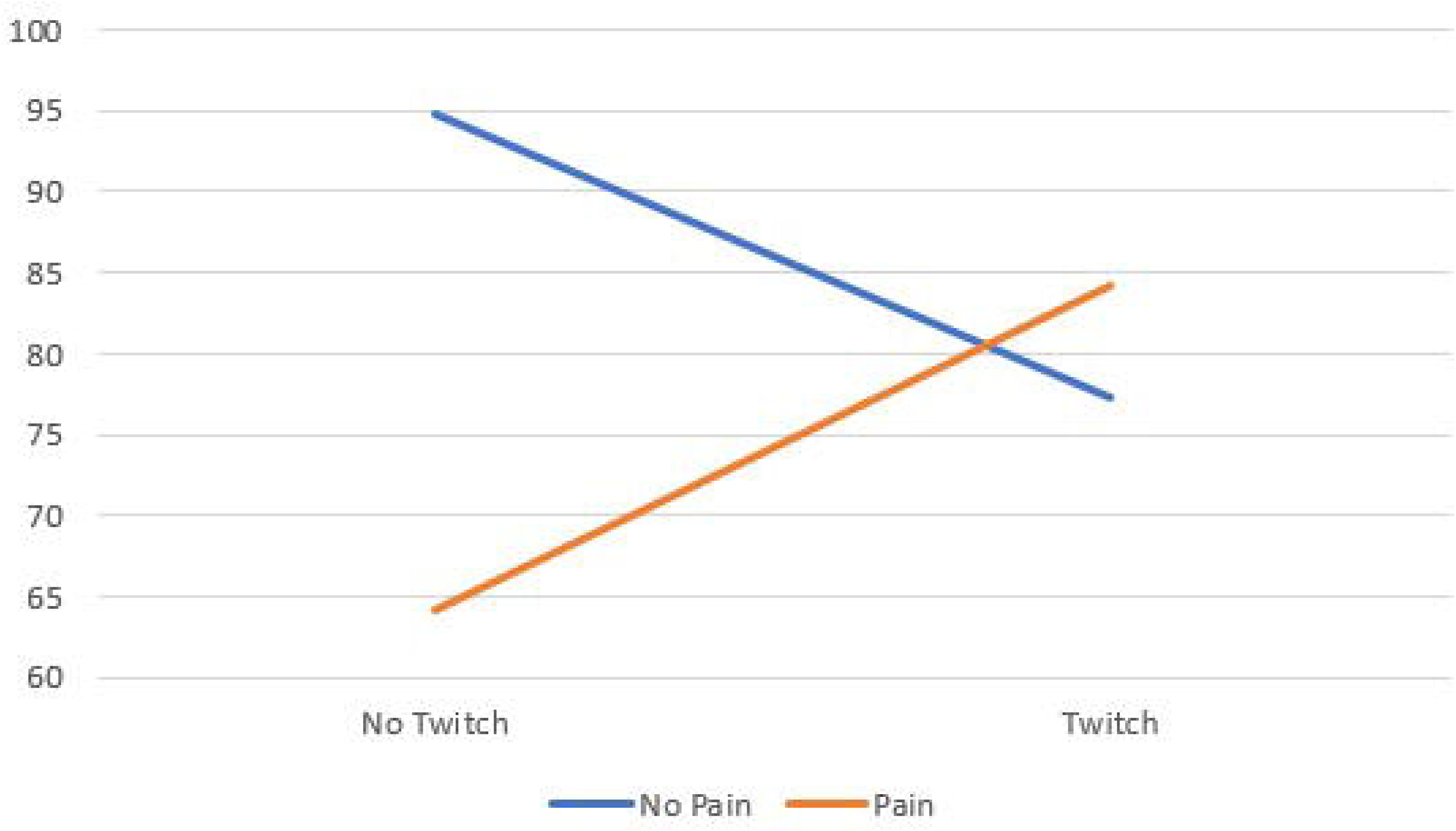
SF-20 Physical Function Subscale.

**Figure 2.**
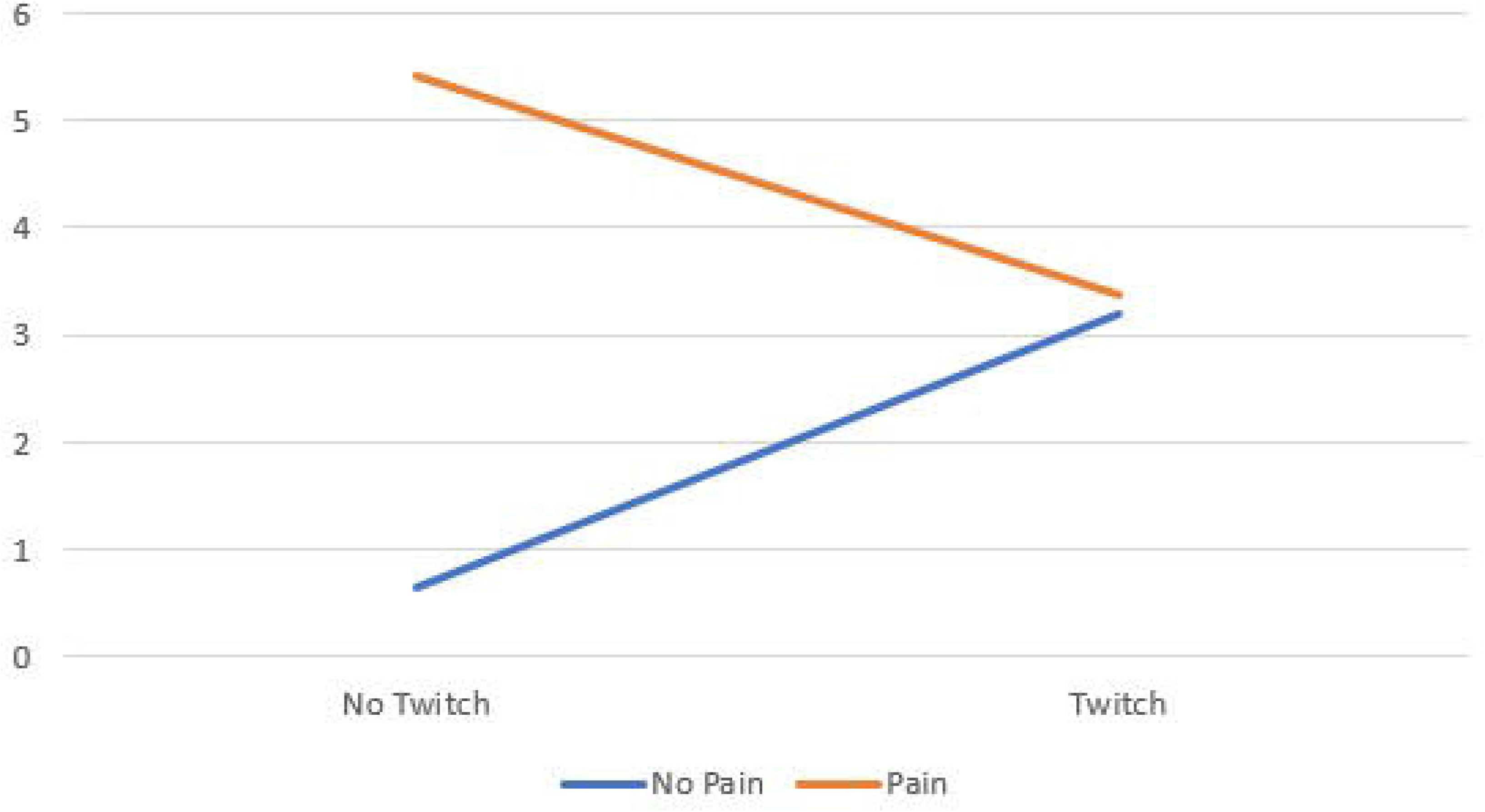
NPR Current Pain Scale.

**Figure 3.**
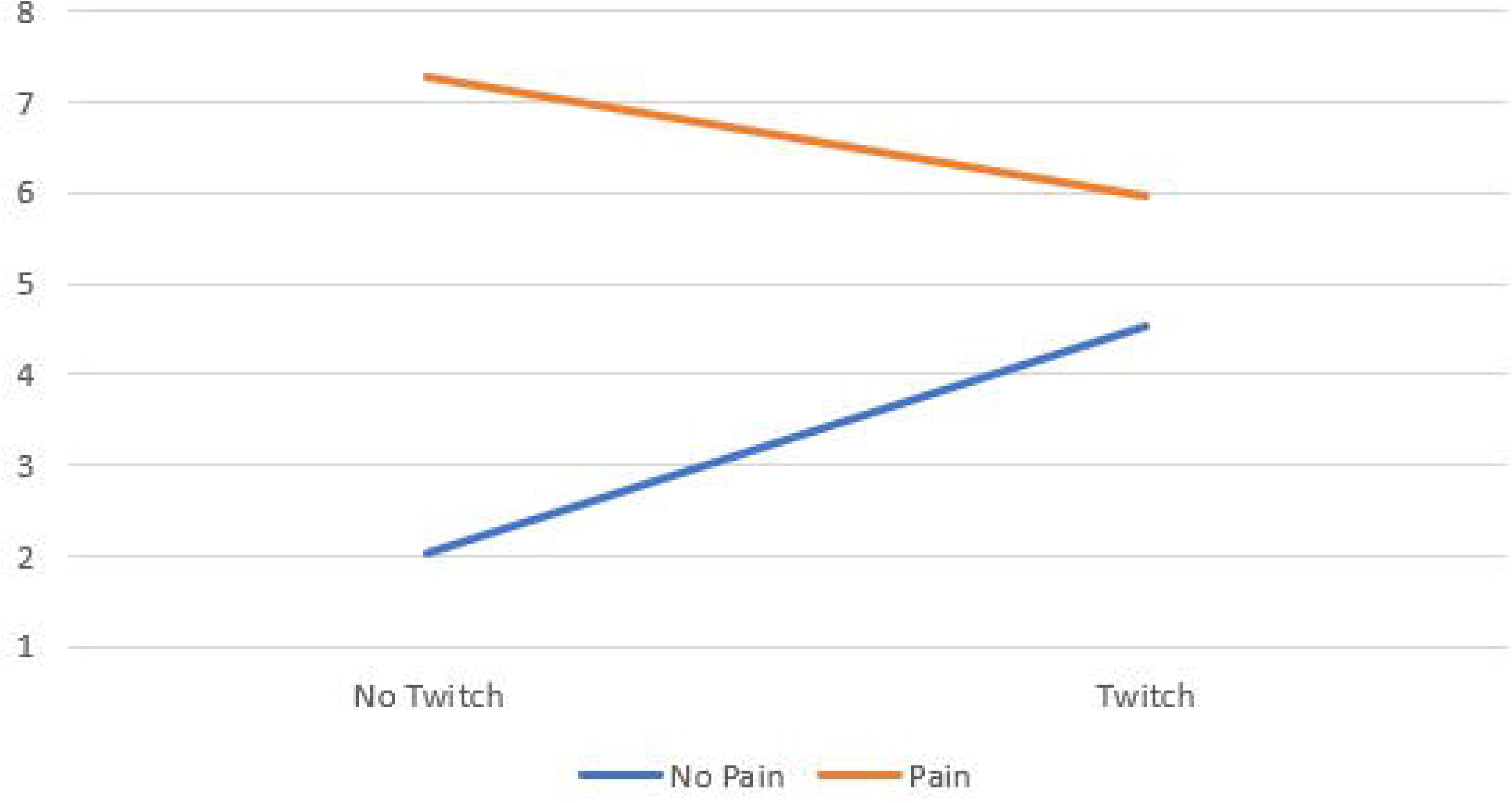
NPR Worst Pain Subscale.

**Table 3.** Clinical Assessment and Questionnaire Intakes by Twitching, Pain, Sex, and Ethnicity Groups.

| Scale | Twitching |  |  |  | Spontaneous Pain |  |  |  | Sex |  |  |  | Ethnicity |  |  |  |
| --- | --- | --- | --- | --- | --- | --- | --- | --- | --- | --- | --- | --- | --- | --- | --- | --- |
|  | Yes (n=18) |  | No (n=7) |  | Yes (n=15) |  | No (n=10) |  | Female (n=18) |  | Male (n=7) |  | Black (n=3) |  | Others (n=22) |  |
|  | M | SD | M | SD | M | SD | M | SD | M | SD | M | SD | M | SD | M | SD |
| MTrPs |  |  |  |  |  |  |  |  |  |  |  |  |  |  |  |  |
| Number of | 4.44 | 0.78 | 4.14 | 1.21 | 4.73 | 0.80 | 3.80 | 0.79 | 4.33 | 0.91 | 4.43 | 0.98 | 4.67 | 0.58 | 4.32 | 0.95 |
| Duration | 77.00 | 80.29 | 97.71 | 103.79 | 57.20 | 71.23 | 121.2 | 94.73 | 109.83 | 87.16 | 13.29 | 6.45 | 84.00 | 60.00 | 82.64 | 89.76 |
| Tenderness (Average) | 4.07 | 1.47 | 3.71 | 1.41 | 4.40 | 1.60 | 3.33 | 0.86 | 3.65 | 0.98 | 4.81 | 2.08 | 5.00 | 2.73 | 3.83 | 1.21 |
| Tenderness (Severe) | 5.50 | 1.50 | 5.29 | 1.50 | 5.80 | 1.52 | 4.90 | 1.29 | 5.22 | 1.31 | 6.00 | 1.83 | 6.67 | 1.53 | 5.27 | 1.42 |
| PPT (Average) | 4.15 | 2.41 | 5.14 | 1.61 | 4.04 | 2.44 | 5.00 | 1.83 | 4.64 | 2.22 | 3.88 | 2.34 | 4.24 | 3.83 | 4.45 | 2.07 |
| PPT (Severe) | 3.36 | 2.03 | 4.34 | 1.34 | 3.31 | 2.13 | 4.11 | 1.44 | 3.86 | 1.81 | 3.05 | 2.13 | 3.46 | 2.83 | 3.65 | 1.82 |
| STS | 10.74 | 2.70 | 10.26 | 1.93 | 10.39 | 2.30 | 10.91 | 2.83 | 10.32 | 2.40 | 11.32 | 2.72 | 14.02 | 1.40 | 10.14 | 2.22 |
| TUG | 9.44 | 1.69 | 9.15 | 2.06 | 9.68 | 1.90 | 8.88 | 1.50 | 9.50 | 1.75 | 9.00 | 1.88 | 11.58 | 1.17 | 9.06 | 1.62 |
| SF-20 |  |  |  |  |  |  |  |  |  |  |  |  |  |  |  |  |
| Physical Function | 81.95 | 19.85 | 86.91 | 17.26 | 82.23 | 17.21 | 85.01 | 22.15 | 77.78 | 19.59 | 97.63 | 4.05 | 72.23 | 25.46 | 84.85 | 18.12 |
| Role Function | 87.50 | 27.45 | 89.29 | 19.67 | 86.67 | 29.68 | 90.00 | 17.48 | 84.72 | 28.62 | 96.43 | 9.45 | 66.67 | 28.87 | 90.91 | 23.84 |
| Social Function | 91.11 | 12.31 | 97.14 | 7.56 | 92.00 | 12.65 | 94.00 | 9.66 | 91.11 | 12.31 | 97.14 | 7.56 | 86.67 | 11.55 | 93.64 | 11.36 |
| Mental Health | 77.01 | 10.40 | 82.29 | 8.28 | 77.33 | 10.55 | 80.21 | 9.33 | 76.12 | 9.70 | 84.57 | 8.46 | 72.70 | 10.97 | 79.27 | 9.85 |
| Health Perceptions | 79.94 | 12.25 | 81.14 | 13.62 | 79.75 | 11.37 | 81.06 | 14.35 | 75.53 | 9.66 | 92.49 | 10.28 | 72.03 | 9.85 | 81.40 | 12.43 |
| Pain | 44.44 | 14.64 | 31.43 | 15.74 | 45.33 | 14.07 | 34.00 | 16.47 | 41.11 | 14.51 | 40.00 | 20.00 | 60.00 | 0.00 | 38.18 | 15.00 |
| ODI | 13.78 | 7.57 | 5.43 | 4.72 | 13.47 | 7.69 | 8.40 | 7.29 | 12.11 | 6.70 | 9.71 | 10.55 | 26.67 | 5.03 | 9.36 | 5.43 |
| PCS | 10.22 | 7.26 | 14.71 | 14.61 | 8.13 | 5.33 | 16.50 | 12.75 | 14.72 | 9.44 | 3.14 | 3.89 | 12.67 | 4.73 | 11.32 | 10.31 |
| PSEQ | 52.33 | 7.11 | 56.57 | 4.20 | 54.40 | 5.01 | 52.20 | 8.66 | 52.39 | 7.09 | 56.43 | 4.47 | 49.67 | 4.73 | 54.05 | 6.76 |
| TSK | 32.72 | 7.05 | 32.29 | 4.42 | 31.80 | 5.80 | 33.80 | 7.22 | 34.17 | 6.01 | 28.57 | 5.65 | 39.00 | 3.61 | 31.73 | 6.17 |
| DASS 21 |  |  |  |  |  |  |  |  |  |  |  |  |  |  |  |  |
| Stress | 7.89 | 5.51 | 8.00 | 10.07 | 8.67 | 5.54 | 6.80 | 8.65 | 8.44 | 7.96 | 6.57 | 2.23 | 9.33 | 1.16 | 7.73 | 7.29 |
| Anxiety | 2.67 | 4.28 | 5.43 | 7.72 | 3.60 | 5.14 | 3.20 | 6.12 | 4.22 | 6.13 | 1.43 | 2.23 | 2.00 | 3.46 | 3.64 | 5.68 |
| Depression | 1.22 | 1.40 | 3.14 | 3.02 | 1.73 | 1.83 | 1.80 | 2.57 | 2.11 | 2.32 | 0.86 | 1.07 | 0.67 | 1.16 | 1.91 | 2.18 |
| EMI |  |  |  |  |  |  |  |  |  |  |  |  |  |  |  |  |
| Stress Management | 3.13 | 1.52 | 3.14 | 2.00 | 3.10 | 1.47 | 3.18 | 1.93 | 2.89 | 1.48 | 3.75 | 1.95 | 1.83 | 0.14 | 3.31 | 1.66 |
| Revitalization | 2.85 | 1.56 | 2.95 | 1.92 | 2.84 | 1.45 | 2.93 | 1.94 | 2.63 | 1.52 | 3.52 | 1.81 | 1.11 | 1.39 | 3.12 | 1.52 |
| Enjoyment | 2.50 | 1.82 | 2.54 | 2.11 | 2.33 | 1.63 | 2.78 | 2.23 | 2.19 | 1.78 | 3.32 | 1.95 | 0.75 | 0.75 | 2.75 | 1.85 |
| Challenge | 2.14 | 1.50 | 1.93 | 1.59 | 2.23 | 1.53 | 1.85 | 1.50 | 1.88 | 1.40 | 2.61 | 1.73 | 0.50 | 0.50 | 2.30 | 1.46 |
| Social Recognition | 1.11 | 1.00 | 0.71 | 0.71 | 0.98 | 1.01 | 1.03 | 0.85 | 0.90 | 0.89 | 1.25 | 1.07 | 0.08 | 0.14 | 1.13 | 0.93 |
| Affiliation | 1.31 | 1.21 | 1.96 | 1.46 | 1.17 | 1.06 | 1.98 | 1.49 | 1.32 | 1.26 | 1.93 | 1.35 | 0.50 | 0.43 | 1.63 | 1.31 |
| Competition | 1.58 | 1.58 | 1.57 | 1.82 | 1.48 | 1.44 | 1.73 | 1.91 | 1.18 | 1.47 | 2.61 | 1.59 | 0.33 | 0.58 | 1.75 | 1.64 |
| Health Pressures | 1.98 | 1.29 | 2.14 | 1.86 | 1.96 | 1.27 | 2.13 | 1.72 | 1.87 | 1.50 | 2.43 | 1.24 | 3.44 | 0.69 | 1.83 | 1.40 |
| Health Avoidance | 3.81 | 0.89 | 3.86 | 1.55 | 3.87 | 0.81 | 3.77 | 1.44 | 3.69 | 1.20 | 4.19 | 0.63 | 4.00 | 0.88 | 3.80 | 1.12 |
| Positive Health | 4.30 | 0.69 | 4.10 | 1.45 | 4.38 | 0.59 | 4.03 | 1.31 | 4.07 | 1.01 | 4.67 | 0.58 | 4.22 | 0.69 | 4.24 | 0.98 |
| Weight Management | 3.74 | 1.48 | 4.64 | 0.28 | 3.68 | 1.55 | 4.45 | 0.72 | 4.10 | 1.30 | 3.71 | 1.45 | 3.33 | 1.44 | 4.08 | 1.31 |
| Appearance | 2.85 | 1.27 | 3.82 | 0.76 | 2.78 | 1.34 | 3.63 | 0.83 | 3.38 | 0.95 | 2.46 | 1.64 | 2.00 | 1.80 | 3.27 | 1.09 |
| Strength & Endurance | 3.86 | 0.97 | 3.14 | 2.00 | 3.83 | 0.89 | 3.40 | 1.83 | 3.42 | 1.43 | 4.29 | 0.81 | 3.92 | 1.04 | 3.63 | 1.38 |
| Nimbleness | 3.26 | 0.92 | 2.86 | 2.07 | 3.22 | 0.97 | 3.03 | 1.75 | 3.07 | 1.32 | 3.33 | 1.36 | 3.11 | 1.02 | 3.15 | 1.36 |
| Numeric Pain Rating |  |  |  |  |  |  |  |  |  |  |  |  |  |  |  |  |
| Current Pain | 3.44 | 1.72 | 1.71 | 2.06 | 3.80 | 1.78 | 1.70 | 1.49 | 2.50 | 1.42 | 4.14 | 2.67 | 4.67 | 2.08 | 2.73 | 1.86 |
| Best Pain past 24 Hr | 1.11 | 1.49 | 0.86 | 1.21 | 1.33 | 1.54 | 0.60 | 1.07 | 1.00 | 1.24 | 1.14 | 1.86 | 4.00 | 1.00 | 0.64 | 0.85 |
| Worst Pain past 24 Hr | 5.78 | 2.16 | 3.00 | 2.58 | 6.27 | 1.71 | 3.10 | 2.51 | 4.72 | 2.35 | 5.71 | 3.15 | 7.67 | 1.53 | 4.64 | 2.48 |

**Table 4.** Factorial ANCOVA Results (*N*=25, *Q[*=1,17)

| Scale | Covariates |  |  |  |  |  |  |  |  |  |  |  | Factors |  |  |  |  |  |  |  |  |
| --- | --- | --- | --- | --- | --- | --- | --- | --- | --- | --- | --- | --- | --- | --- | --- | --- | --- | --- | --- | --- | --- |
|  | Age |  |  | Gender |  |  | Asian |  |  | Black |  |  | Twitching |  |  | Spontaneous Pain |  |  | Twitch*Pain |  |  |
| | F | p | $\eta^2$ | F | p | $\eta^2$ | F | p | $\eta^2$ | F | p | $\eta^2$ | F | p | $\eta^2$ | F | p | $\eta^2$ | F | p | $\eta^2$ |
| **MTrPs** |  |  |  |  |  |  |  |  |  |  |  |  |  |  |  |  |  |  |  |  |  |
| Number | 1.06 | .32 | .06 | 1.23 | .28 | .07 | 0.64 | .43 | .04 | 0.11 | .75 | .01 | 0.14 | .72 | .01 | **11.11** | **.004\*\*** | **.40** | 3.66 | .07 | .18 |
| Duration | 1.05 | .32 | .06 | **4.48** | **.05\*** | **.21** | 3.64 | .07 | .18 | 0.09 | .77 | .01 | 0.87 | .37 | .05 | **5.93** | **.03\*** | **.26** | 1.94 | .18 | .10 |
| Tenderness (Average) | 0.09 | .76 | .01 | 3.23 | .09 | .16 | 0.04 | .85 | .002 | 1.54 | .23 | .08 | 0.28 | .61 | .02 | 4.08 | .06 | .19 | 1.70 | .21 | .09 |
| Tenderness (Severe) | 0.04 | .85 | .002 | 1.29 | .27 | .07 | 0.07 | .79 | .004 | 2.00 | .18 | .11 | 0.40 | .54 | .02 | 2.76 | .12 | .14 | 1.00 | .33 | .06 |
| PPT(Average) | 0.49 | .50 | .03 | 1.73 | .21 | .09 | 1.82 | .20 | .10 | 0.09 | .76 | .01 | 0.58 | .46 | .03 | 0.97 | .34 | .05 | 1.60 | .22 | .09 |
| PPT Severe | 1.19 | .29 | .07 | 2.09 | .17 | .11 | 2.45 | .14 | .13 | 0.19 | .67 | .01 | 1.04 | .32 | .06 | 0.65 | .43 | .04 | 1.27 | .28 | .07 |
| STS | 0.42 | .53 | .02 | 1.11 | .31 | .06 | 0.07 | .80 | .004 | **6.48** | **.02\*** | **.28** | 0.03 | .86 | .002 | 0.03 | .86 | .002 | 2.07 | .17 | .11 |
| TUG | 3.58 | .07 | .17 | 0.01 | .93 | <.001 | 0.26 | .62 | .02 | 3.20 | .09 | .16 | 0.83 | .38 | .05 | 3.90 | .07 | .19 | 2.09 | .17 | .11 |
| **SF-20** |  |  |  |  |  |  |  |  |  |  |  |  |  |  |  |  |  |  |  |  |  |
| Physical Function | 3.02 | .10 | .15 | 2.04 | .17 | .11 | 0.12 | .73 | .01 | 0.25 | .62 | .02 | 0.03 | .87 | .002 | 2.39 | .14 | .12 | **5.10** | **.04\*** | **.23** |
| Role Function | 4.35 | .02 | .20 | 0.01 | .92 | .001 | 0.63 | .44 | .04 | 1.34 | .26 | .07 | 0.01 | .92 | .001 | 0.79 | .39 | .05 | 0.96 | .34 | .05 |
| Social Function | 1.35 | .26 | .07 | 0.41 | .53 | .02 | 0.54 | .47 | .03 | 0.03 | .87 | .002 | 0.28 | .61 | .02 | 0.32 | .58 | .02 | 0.92 | .35 | .05 |
| Mental Health | 0.59 | .46 | .03 | 4.31 | .05 | .20 | 0.41 | .53 | .02 | 1.58 | .23 | .09 | 0.69 | .42 | .04 | 0.02 | .88 | .001 | 0.99 | .33 | .06 |
| Health Perceptions | 0.003 | .96 | <.001 | **7.30** | **.02\*** | **.30** | 2.03 | .17 | .11 | 3.32 | .09 | .16 | 0.06 | .81 | .004 | 1.67 | .21 | .09 | 1.18 | .29 | .07 |
| Pain | 1.68 | .21 | .09 | <.001 | 1.00 | <.001 | 0.55 | .47 | .03 | **6.43** | **.02\*** | **.27** | 0.43 | .52 | .03 | 4.10 | .06 | .03 | 2.69 | .12 | .14 |
| ODI | 1.58 | .23 | .09 | 0.79 | .39 | .04 | 0.06 | .80 | .004 | **19.35** | **<.001\*\*\*** | **.53** | 2.88 | .11 | .15 | 3.15 | .09 | .16 | 0.47 | .50 | .03 |
| PCS | 0.09 | .77 | .01 | **6.76** | **.02\*** | **.29** | 3.12 | .10 | .16 | 1.14 | .30 | .06 | 0.01 | .92 | .001 | **5.55** | **.03\*** | **.25** | 2.07 | .17 | .11 |
| PSEQ | 0.01 | .91 | .001 | 0.71 | .41 | .04 | 0.12 | .73 | .01 | 0.57 | .46 | .03 | 2.51 | .13 | .13 | 1.23 | .28 | .07 | 0.01 | .92 | .001 |
| TSK | 0.10 | .76 | .01 | 1.17 | .30 | .06 | 0.45 | .51 | .03 | 3.54 | .08 | .17 | 0.001 | .98 | <.001 | 0.001 | .97 | <.001 | 1.54 | .23 | .08 |
| **DASS 21** |  |  |  |  |  |  |  |  |  |  |  |  |  |  |  |  |  |  |  |  |  |
| Stress | 0.09 | .77 | .01 | 1.76 | .20 | .09 | 0.10 | .76 | .01 | 0.13 | .72 | .01 | 0.05 | .83 | .003 | <0.001 | 1.00 | <.001 | 3.75 | .07 | .181 |
| Anxiety | 0.27 | .61 | .02 | 2.49 | .13 | .13 | 1.17 | .29 | .07 | 0.17 | .68 | .01 | 1.95 | .18 | .10 | 0.29 | .60 | .02 | 0.48 | .50 | .03 |
| Depression | 1.33 | .27 | .07 | 2.38 | .14 | .12 | 0.04 | .84 | .002 | 0.52 | .48 | .03 | 4.04 | .06 | .19 | 0.27 | .61 | .02 | 1.44 | .25 | .08 |
| **EMI** |  |  |  |  |  |  |  |  |  |  |  |  |  |  |  |  |  |  |  |  |  |
| Stress Management | 0.59 | .45 | .03 | 2.47 | .13 | .13 | 0.26 | .62 | .02 | 3.17 | .09 | .16 | 0.04 | .85 | .002 | 0.04 | .86 | .002 | 1.70 | .21 | .09 |
| Revitalization | 0.18 | .67 | .01 | 1.54 | .23 | .08 | 1.13 | .30 | .06 | **5.36** | **.03\*** | **.24** | 0.01 | .94 | <.001 | 0.003 | .96 | <.001 | 0.90 | .36 | .05 |
| Enjoyment | 0.01 | .94 | <.001 | 1.45 | .24 | .08 | 1.02 | .33 | .06 | 3.78 | .07 | .18 | 0.10 | .76 | .01 | 0.31 | .58 | .02 | 0.48 | .50 | .03 |
| Challenge | 2.14 | .16 | .11 | 0.26 | .62 | .02 | 0.05 | .83 | .003 | 2.58 | .13 | .13 | 0.15 | .70 | .01 | 0.09 | .77 | .01 | <.001 | 1.00 | <.001 |
| Social Recognition | 1.64 | .22 | .09 | 0.84 | .37 | .05 | 0.48 | .50 | .03 | 2.35 | .14 | .12 | 2.70 | .12 | .14 | 0.47 | .50 | .03 | 0.05 | .83 | .003 |
| Affiliation | 0.23 | .64 | .01 | 3.52 | .08 | .17 | 0.04 | .84 | .002 | 2.32 | .15 | .12 | 0.18 | .67 | .01 | 0.46 | .51 | .03 | 3.09 | .10 | .15 |
| Competition | 3.37 | .08 | .17 | 3.86 | .07 | .19 | 0.92 | .35 | .05 | 0.76 | .40 | .04 | 0.57 | .46 | .03 | 0.56 | .46 | .03 | 0.05 | .82 | .003 |
| Health Pressures | 2.79 | .11 | .14 | 2.30 | .15 | .12 | 0.03 | .87 | .002 | 1.35 | .26 | .07 | 0.23 | .64 | .01 | 0.01 | .92 | .001 | 0.93 | .35 | .05 |
| Health Avoidance | 1.50 | .24 | .08 | 1.54 | .23 | .08 | 0.65 | .43 | .04 | 0.12 | .74 | .01 | 0.18 | .68 | .01 | 0.36 | .56 | .02 | 1.35 | .26 | .07 |
| Positive Health | 0.25 | .62 | .01 | 2.60 | .13 | .13 | 0.23 | .64 | .01 | 0.23 | .64 | .01 | 0.01 | .94 | <.001 | 1.33 | .26 | .07 | 2.68 | .12 | .14 |
| Weight Management | 1.07 | .32 | .06 | 0.71 | .41 | .04 | 0.06 | .81 | .004 | 0.05 | .82 | .003 | 1.16 | .30 | .07 | 0.46 | .51 | .03 | <.001 | 1.00 | <.001 |
| Appearance | 0.62 | .44 | .04 | 4.18 | .06 | .20 | 0.11 | .75 | .01 | 1.08 | .31 | .06 | 1.30 | .27 | .07 | 1.34 | .26 | .07 | 0.62 | .44 | .04 |
| Strength & Endurance | 0.02 | .89 | .001 | 1.24 | .28 | .07 | 0.02 | .89 | .001 | 0.001 | .98 | <.001 | 0.67 | .43 | .04 | 0.04 | .85 | .002 | 0.06 | .82 | .003 |
| Nimbleness | 0.08 | .78 | .01 | 0.13 | .73 | .01 | 0.38 | .55 | .02 | 0.16 | .70 | .01 | 0.10 | .75 | .01 | 0.06 | .81 | .004 | 0.29 | .60 | .02 |
| **Numeric Pain Rating** |  |  |  |  |  |  |  |  |  |  |  |  |  |  |  |  |  |  |  |  |  |
| Current Pain | 0.21 | .65 | .01 | **8.92** | **.01** | **.34** | 1.77 | .20 | .09 | 1.54 | .23 | .08 | 0.17 | .68 | .01 | **17.83** | **<.001\*\*\*** | **.51** | **13.03** | **.002\*\*** | **.43** |
| Best Pain | 1.34 | .26 | .07 | 0.00 | .98 | <.001 | 1.76 | .20 | .09 | **33.05** | **<.001\*\*\*** | **.66** | **4.98** | **.04** | **.23** | **8.45** | **.01\*** | **.33** | 1.79 | .20 | .10 |
| Worst Pain | 0.14 | .72 | .01 | 0.46 | .51 | .03 | 1.74 | .21 | .09 | 3.56 | .08 | .17 | 0.48 | .50 | .03 | **17.10** | **<.001\*\*\*** | **.50** | **4.79** | **.04\*** | **.22** |

Participants with twitching responses reported a higher level of best pain score in the past 24 hours compared to those who did not have twitching responses, with large effect size (*F*[1,17]=4.98, *p*=.04, η^2^=.23; *M* =1.11±1.49 vs. (*M*=0.86±1.21). This indicates that participants with active and latent MTrPs report higher levels of best pain (i.e., their “best pain” consisted of higher pain levels) as compared to those with atypical MTrPs. Further, participants in groups which reported pain (Groups 1 and 3) also had more trigger points (*F*[1,17]=11.11, *p*=.004, η^2^=.40; *M*=4.73±0.80 vs. *M*=3.80±0.79); shorter period of pain history (*F*[1,17]=5.93, *p*=.03, η^2^=.26; *M*=57.20±71.23 vs. *M*=121.20±94.73); low PCS total scores (*F*[1,17]=5.55, *p*=.03, η^2^=.25; *M*=8.13±5.33 vs. *M*=16.50±12.75); higher level of current pain (*F*[1,17]=17.83, *p*<.001, η^2^=.51; *M*=3.80±1.78 vs. *M*=1.70±1.49); best pain level in the past 24 hours (*F*[1,17]=8.45, *p*=.01, η^2^=.33; *M*=1.33±1.54 vs. *M*=0.60±1.07); and worst pain level in the past 24 hours (*F*[1,17]=17.10, *p*<.001, η^2^=.50; *M*=6.27±1.71 vs. *M*=3.10±2.51) compared to those who were in groups that did not report being in pain (Group 2 and 4) with a large effect size. That is, regardless of twitching response, participants with spontaneous pain had more MTrPs, shorter pain history, higher pain catastrophizing score, and higher current pain, best pain and worst pain when compared to those who reported no pain.

Female participants had a longer history of pain (*F*[1,17]=4.48, *p=*.05, η^2^=.21; *M*=109.83±87.16 vs. *M*=13.29±6.45), lower level of health perceptions (*F*[1,17]=7.30, *p=*.02, η^2^=.30; *M*=75.53±9.66 vs. *M*=92.49±10.28), higher PCS score (*F*[1.17]=6.76,*p=*.02, η2= .29; *M*=14.72±9.44 vs. *M*=3.14±3.89), and lower level of current pain (*F*[1.17]=8.92, *p=*.01, η^2^=.34; *M*=2.50±1.42 vs. *M*=4.14±2.67) than male participants (Table 3 and Table 4). In addition, Black participants had worse STS score (*F*[1,17]=6.48, *p*=.02, η^2^=.28; *M*=14.02±1.40 vs. *M*=10.14±2.22), higher pain component in QoL (*F*[1,17]=6.43, *p=*.02, η^2^=.27; *M*=60.00±0.00 vs. *M*=38.18±15.00), higher ODI scores (*F*[1,17]=19.35, *p*<.001, η^2^=.53; *M*=26.67±5.03 vs. *M*=9.36±5.43), lower EMI revitalization scores (*F*[1,17]=5.36, *p=*.03, η^2^=.24; *M*=1.11±1.39 vs. *M*SD=3.12±1.52), and higher levels of best pain in the past 24 hours (*F*[1,17]=33.05, *p*<.001, η^2^=.66; *M*=4.00±1.00 vs. *M*=0.64±0.85) compared with other races. Identifying as Asian and the age of the participants had no impact on each measure.

### Treatment or strategies used to treat MPS

Popular type of treatment or strategies for MPS included eating a healthy diet (*n*=22), exercising (*n*=22), stretching (*n*=22) and use of a pain pill (*n*=20), followed by using heat and ice (*n*=17), use of relaxation techniques (*n*=17), massage therapy (*n*=14), improving sleep quality (*n*=12), practicing yoga (*n*=10), physical therapy (*n*=9), chiropractic care (*n*=8), meditation (*n*=7), posture training (*n*=7) and osteopathic manipulation (*n*=6) (Table 5). Professional help strategies would include physical therapy, chiropractic care, massage therapy, and osteopathic manipulation treatments, while the rest would be considered as self-help strategies. Among the total of 212 strategies chosen to treat MPS by participants, 77% (*n*=164) were self-care and 23% were professional help (*n*=48). The top six treatments/strategies chosen included eating a healthy diet, exercising, stretching, pain pills (mostly over the counter), heat and ice, and relaxation techniques are considered self-care.

**Table 5.** Treatment or Strategies Used to Manage MPS.

| Treatment and strategy | Self-care | Professional help | N | % |
| --- | --- | --- | --- | --- |
| Eating healthy diet | X |  | 22 | 10.38 |
| Exercising | X |  | 22 | 10.38 |
| Use of stretching | X |  | 22 | 10.38 |
| Use of pain medication | X |  | 20 | 9.43 |
| Heat/cold | X |  | 17 | 8.02 |
| Practice relaxation | X |  | 17 | 8.02 |
| Use of Massage |  | X | 14 | 6.60 |
| Enough sleep | X |  | 12 | 5.66 |
| Practicing yoga | X |  | 10 | 4.72 |
| Use of physical therapy |  | X | 9 | 4.25 |
| Seeing a chiropractor |  | X | 8 | 3.77 |
| Practice meditation | X |  | 7 | 3.30 |
| Posture training | X |  | 7 | 3.30 |
| Use of osteopathic manipulation |  | X | 6 | 2.83 |
| Use of ultrasound |  | X | 3 | 1.42 |
| Use of TENS unit |  | X | 3 | 1.42 |
| Use of Injections |  | X | 2 | 0.94 |
| Use of acupuncture |  | X | 2 | 0.94 |
| Use of back brace | X |  | 2 | 0.94 |
| Talking to a counselor |  | X | 1 | 0.47 |
| Going to a support group | X |  | 1 | 0.47 |
| Use of cream | X |  | 1 | 0.47 |
| Dog hugging | X |  | 1 | 0.47 |
| Drinking water | X |  | 1 | 0.47 |
| Watching soap opera/news | X |  | 1 | 0.47 |
| Working in garden | X |  | 1 | 0.47 |
| N (%) | 164 (77%) | 48 (23%) | 212 | 100 |

## Discussion

This is one of few studies investigating characteristics of patients with MPS of the low back. This study identified several findings that could potentially assist in understanding MPS of the low back. A similar study with a non-US population revealed 63.5%-90% of patients with low back pain have MPS.^9, 10^ Likewise, the current study found that 100% of patients show more than one visible nodule on the low back muscle as identified by an ultrasound procedure, and 72% of individuals met the criteria of having active or latent MPS

The current study found that not all nodules met the definitions provided by Travell and Simons.^3^ There are a couple of potential explanations: First, several past studies have pointed out that manual palpation for identifying myofascial trigger points is not as reliable as what we would expect for a diagnostic option.^45, 46^ For example, one of the most recent studies shows the inter-rater reliability is between 0.49 and 0.75 for low leg muscles.^46^ Potentially, patients with atypical symptoms could be misdiagnosed due to poor reliability of palpation. Second, it is possible that atypical MTrPs may be in the process of becoming active or latent MTrPs or dissolving as the disease process is ending in that area and returning to normal. Alternatively, one study proposed additional diagnostic criteria^47^ which postulated that atypical MTrPs are also a sign of MPS. Finally, a systematic review study found that in clinical trials, the MTrPs diagnostic criteria utilized were widely different. However, the most popular combination is spot tenderness, referred pain, and local twitch response.^48^ With the current level of understanding of MPS none of these possibilities can be ruled out. Additional studies are needed, including finding objective ways to identify MTrPs and define MPS.

This study found multiple MTrPs and mixed numbers of MTrPs, which is similar to findings in other studies examining patients with low back pain, as well as studies of patients with fibromyalgia.^12, 49^ One group of non-specific low back pain patients and a matching control group found patients with non-specific low back pain had an average of 5.5 mixed MTrPs.^12^ In a patient population with fibromyalgia, multiple trigger points seem to relate to having a pain complaint. Women with fibromyalgia had an average of 11 MTrPs, with the majority of them active MTrPs, while healthy control patients only had latent MTrPs with an average of two MTrPs.^49^ It is unclear whether MPS is a unique symptom with a specific musculoskeletal disorder or a common syndrome of several musculoskeletal pain conditions. Findings in current study seems to support the latter.

Participants with pain reports had more MTrPs compared to those who reported no pain. A study with low back pain patients also found the number of active MTrPs was associated with pain intensity.^12^ This suggests the number of MTrPs or number of active MTrPs, instead of other MPS characteristics, may be the most important predictors for the level of pain complaints in MPS patients. In addition, complaints of pain seem to be a decisive factor associated with worse physical function, but twitching response is not. This study found that in addition to having more MTrPs, participants who reported pain had shorter pain histories, higher pain catastrophizing scores, and higher pain report levels compared to those who reported no pain. The causal relationship between pain reports and catastrophizing in this crossover study cannot be determined. However, past studies have revealed that a higher level of pain catastrophizing was associated with developing higher pain intensity and higher levels of disability in patients with temporomandibular dysfunction, a condition commonly caused by MPS.^50^

Participants in the current study were a relatively healthy young to middle-age adult population. However, approximately 80% of participants took pain medications. One study working with middle age patients found that more than 60% used pain medication for their low back pain.^51^ Our study population is younger and reported lower levels of pain, yet a higher percentage of them used pain medications compared to the previously cited study population. Similar trends were also observed in an international population with musculoskeletal pain.^52^ Potential reasons could be that the younger generations are busier, seeking instant relief of pain, and/or have less financial means for alternative therapies. If this is true, this may have some clinical implications. MPS cannot be cured simply with pharmacological intervention. Without life-style changes, patients’ level of MPS may increase in terms of severity and chronicity, resulting in poor quality of life and even leading to future opioid abuse.

We cannot find articles examining the treatment choices of patients with MPS of the low back. This study found that in addition to the use of pain pills (mostly OCT), the top 5 treatments/strategies chosen were in the category of self-care. Indeed, among all strategies chosen to treat MPS, 77% were self-care. This is similar to a common clinical practice that recommends self-care strategies, including improving level of physical activity, exercising, posture, and reduction of stress.^53^ These self-care behaviors were also reported by patients with myofascial temporomandibular disorder pain.^54^

Female participants had longer history of pain, higher catastrophizing scores, and lower levels of current pain than male participants. This study only included a few Black participants. However, we found that black participants had worse physical function, higher level of disability, and higher level of best pain in the past 24 hours compared with other races. Again, no studies were found for an MPS population, but in patients with low back pain, similar findings were reported in which Black participants also reported higher level of pain severity^51, 55^ and higher levels of disability compared to White participants, but the study did not find a gender difference in pain reporting.^55, 56^

Limitations of this study include a small sample size; thus, findings should be considered cautiously. In addition, the cross-sectional design of this study limits its ability to make causal inferences. Finally, this is a highly educated group of participants, therefore the results cannot be generalized to other populations.

## Author Contributions

PT, JLE, CW, and JE designed the study. PT, JLE, CW, MWG, and KM carried out the study and analyzed MTrP and clinical data. All authors contributed to the writing of results from this study.

## Author Declarations

The authors do not declare any conflicts of interest in the present study.

## Data Availability

All data produced in the present study are available upon reasonable request to the authors.

## Acknowledgements

This study was supported by the Research Eureka Accelerator Program (REAP), Edward Via College of Osteopathic Medicine.

